# Pulsed electromagnetic fields may be effective for the management of primary osteoporosis: a systematic review and meta-analysis

**DOI:** 10.1101/2021.05.30.21258065

**Authors:** Siyi Zhu, Yi Li, Liqiong Wang, Jinming Huang, Kangping Song, Xinling Gan, Xiaona Xiang, Chengqi He, Lin Yang

**Affiliations:** Rehabilitation Medical Center, West China Hospital, Sichuan University, Chengdu, Sichuan, China; Rehabilitation Medicine key laboratory of Sichuan Province, West China Hospital, Sichuan University, Chengdu, Sichuan, China

**Author notes:** Corresponding author: Dr. Chengqi He or Dr. Qiang Gao or Dr. Lin Yang at the Rehabilitation Medical Center, West China Hospital, Sichuan University, No. 37 Guoxue Alley, Wuhou District, 610041, Chengdu, China. (CH) or (LY).

**Keywords:** Osteoporosis, Electromagnetic Fields, Bone Density, Physical Therapy Modalities, Disability Evaluation

## Abstract

**Objective:** To investigate the effectiveness of pulsed electromagnetic fields (PEMFs) for the management of primary osteoporosis in older adults.

**Design:** Systematic review and meta-analysis.

**Data Sources:** MEDLINE, EMBASE, Web of Science, CENTRAL and CCTR, Physiotherapy Evidence Database, CNKI, VIP, Wan Fang, ClinicalTrials.gov and Current controlled trials from the inception dates to April 30, 2021.

**Eligibility criteria for study selection:** Randomised controlled trials or quasi-randomised trials examining the effects of PEMFs compared to placebo or sham or other agents for the management of primary osteoporosis (including those with previous fractures).

**Data extraction and synthesis:** Two independent reviewers extracted data. Primary outcomes were bone mass and number of incident fractures. Secondary outcomes were functional assessments, quality of life, and adverse events. Risk of bias was assessed with the Cochrane Collaboration’s tool and certainty of evidence with the grading of recommendations assessment, development and evaluation (GRADE) framework. A random effects model was used to calculate mean differences and 95% confidence intervals.

**Results:** Eight trials including 396 participants met the inclusion criteria. Low certainty evidence showed that PEMFs was non-inferior to conventional pharmacological agents in preventing the decline of Bone Mineral Density (BMD) at the lumbar (MD 0.01; CI -0.04 to 0.06) and femur neck (MD 0.01; CI -0.02 to 0.04), and improving balance function measured by Berg Balance Scale (BBS) (MD 0.01; CI -0.09 to 0.11) and Timed Up and Go test (MD -0.04; CI -0.80 to 0.72), directly after intervention. The similar effects were observed in BMD and BBS at 12- and 24-weeks follow-up from baseline with moderate certainty evidence. Very low certainty evidence showed that PEMFs (versus exercise) had small but significant effect on BMD at the femur neck (MD 0.10; CI 0.01 to 0.20), and no effect on BMD at the lumbar (MD 0.15; CI -0.04 to 0.35).

**Conclusion:** PEMFs had positive effects non-inferior to first-line treatment on BMD and balance function in older adults with primary osteoporosis, but with low to very low certainty evidence and short-term follow-ups. There is a need for high-quality randomised controlled trials evaluating PEMFs for the management of primary osteoporosis.

**Registration:** PROSPERO CRD42018099518.

## INTRODUCTION

Osteoporosis is a systemic and multifactorial skeletal disorder characterized by low bone mineral density and skeletal fragility that occur with aging, with a consequent increase of susceptibility to low-trauma fractures ^1^. The most prevalent symptoms of osteoporosis are fractures at vertebrae, proximal femur (hip), and wrist affecting patients’ physical function and quality of life. It is estimated that over 200 million people worldwide are affected by osteoporosis, accounting for 8.9 million fractures annually ^2^. The possibility of osteoporotic fractures exceeds 40% and the probability of hip fracture alone could target 20% in white female population over 50 years old ^3^. In China, a higher incidence of hip fractures in men than in women was reported ^4^. Each year, osteoporotic fractures account for over 432,000 hospitalizations and 2.5 million medical visits in the USA ^5^. The treatment costs for fractures were recorded at nearly $17 billion in the USA in 2005 ^6^, and €31.7 billion in Europe in 2000 ^7^. Therefore, osteoporosis has been identified as a major health burden globally by WHO, due to its high prevalence, disability rate, related mortality and poor quality of life ^8^.

Rehabilitation interventions given its important roles in modifying risk factors related to fractures, restoring function and improving quality of life are frequently recommended as an option in the nonpharmacological management of osteoporosis ^9,10^. Pulsed electromagnetic fields (PEMFs) at a specific intensity and frequency have been proved effective in attenuating bone loss and the relief of pain and discomfort after osteoporosis. As we reviewed elsewhere ^11^, PEMFs were found to be positive at promoting bone formation by stimulating the formation and differentiation of osteoblasts, and negative at inhibiting the function of osteoclasts in bone resorption. Experimental studies suggest that PEMFs may exert effects on Ca^2+^-related receptors on the bone cell membrane which play a regulatory role in the maintenance of bone remodelling ^12^. Further, the exposure of PEMFs could influence the physiopathology of osteoporosis by targeting inflammation and potentially relieving pain via these regulatory processes and improvements in bone remodeling ^13^.

PEMFs have been widely used as an clinical option for the management of pain and discomfort related to osteoporosis since the introduce of its usage for non-union fractures was approved by FDA in 1979 ^14^. However, clinical trials evaluating the effectiveness of PEMFs have been conducted with inconsistent results, to which parameters of PEMFs used in studies, follow-up time points and clinical settings differ across studies may lead ^11^. In order to expand upon the current knowledge on whether PEMFs is an effective physical agent for osteoporosis clinically, a systematic review and meta-analysis of clinical trials was performed to compare PEMFs with placebo or sham or other agents for the management of primary osteoporosis in older adults.

## METHODS

This systematic review and meta-analysis was conducted in accordance with the Cochrane Handbook for Systematic Reviews of Interventions ^15^and reported based on Preferred Reporting Items for Systematic Reviews and Meta-analyses guidelines (PRISMA) ^16^. The protocol of this study is available in PROSPERO (CRD42018099518) ^17^.

### Identification and selection of studies

We searched the MEDLINE (via Ovid), EMBASE (via Ovid), Web of Science, CENTRAL and CCTR (via The Cochrane Library), Physiotherapy Evidence Database (via PEDro website), CNKI, VIP, Wan Fang, ClinicalTrials.gov (https://www.clinicaltrials.gov/) and Current controlled trials (www.controlled-trials.com) from the inception dates to December 9, 2018, using the keywords *pulsed electromagnetic fields* and *osteoporosis*. The Open Grey (http://www.opengrey.eu/) was searched for the Grey Literature research. The detailed electronic search strategies are provided in Supplementary Appendix I. An additional search was performed under a mechanism of living systematic review ^18^to identify recently published randomized clinical trials (RCTs) from December 10, 2018 to April 30, 2021 using the databases and keywords described above. The whole procedure was assisted by a librarian from Sichuan University.

Randomised controlled trials or quasi-randomised trials examining the effects of PEMFs compared to placebo or sham or other agents for the management of primary osteoporosis (including those with previous fractures) were included if they met the inclusion criteria listed in Box 1. Studies were excluded if the study population had a diagnosis of corticosteroid-induced osteoporosis or other secondary osteoporosis (e.g., rheumatoid arthritis), studies where participants had a history of hip replacement or surgery related with osteoporotic fractures, the study type was observational studies, review articles, abstracts, conference reports and book chapters.

A three-stage screening methodology was performed to select relevant RCTs for this review. Primarily, all titles were screened by one reviewer (SYZ) for eligibility and irrelevant papers were excluded accordingly. Secondary, two reviewers (YL and LQW or KPS) independently reviewed each study title and abstract. Thirdly, two independent reviewers (XNX and JMH or XLG) accessed the full text to assess against the eligibility criteria for each potentially eligible study. A third reviewer (CQH or LY) was involved for any disagreement.

### Assessment of characteristics of studies

#### Quality assessment

The risk of bias was assessed by using the Cochrane Collaboration’s ‘Risk of bias’ tool ^15^. Seven key domains were assessed by two reviewers (SYZ and LQW): 1) the randomization sequence generation, 2) allocation concealment, 3) blinding of participants and personnel, 4) blinding of outcome assessment, 5) incomplete outcome data, 6) selective reporting, and 7) other bias. The included studies were graded as low, unclear, or a high risk of bias. Methodological quality was assessed with the use of Physiotherapy Evidence Database (PEDro) tool ^19^, which was proved reliable ^20^ and valid ^21^. Each criterion in the PEDro scale with a range of 0-10 was scored 1 (“yes”) or 0 (“no, don’t know/unclear”). Generally, trials with a PEDro summary score of over five. were considered to have adequate methodological quality ^22^. Finally, we used Grading of Recommendations, Assessment, Development and Evaluations (GRADE) ^23^ to describe the overall quality of the body of evidence.

#### Participants

To be included, studies involved participants were healthy older adults (including those with previous fractures) aged over 50 years with primary osteoporosis ^24^, recognized by two distinct types ^25^: 1) type I occurred in postmenopausal women; 2) type II, known as senile osteoporosis, occurred in both men and women.

#### Interventions

All RCTs applying electromagnetic fields with pulsed signal and extremely low frequencies (between 5 and 300 Hz) for the management of primary osteoporosis were included. The parameters (frequency and intensity) of PEMFs, sessions per week and total duration of the treatment period were recorded to describe the interventions.

#### Outcome measures

All outcomes were continuous data and recorded as the percent change from baseline to post-intervention and different follow-up timepoints. To be included, trials had to provide original data or sufficient information about at least one of outcomes on bone mass, number of incident fractures, self-reported data on the changes in balance and quality of life, physical activity and function, and adverse events. Primary outcomes were bone mass (e.g., Bone Marrow Density or Bone Mineral Content) immediately post-intervention and at follow-ups, and number of incident fractures. Secondary outcomes were functional assessments (e.g., Berg Balance Scale, Timed Up and Go test), quality of life (e.g., EuroQoL (EQ 5D)) and adverse events (e.g., falls and death).

### Data extraction and analysis

Two independent reviewers (SYZ and YL) extracted the following information from eligible studies: lead author; year of publication; original country; subject characteristics; study design; treatment information; intervention protocol; outcome measures; raw outcome data; follow-up period and other relevant information. Disagreements were resolved by consensus.

All meta-analyses were performed using analysis was performed using Review Manager (RevMan) software (The Cochrane Collaboration, version 5.4). For each included study, the mean difference (MD) of percentage change with 95% confidential intervals (CIs) was calculated when the outcome measures were consistent across studies or else the standard mean difference (SMD) was calculated instead for continuous outcomes (reporting mean and standard deviation (SD) or standard error (SE) of the mean). If the MD was not reported, it was calculated as the change between values of the baseline and post-intervention. In the case that the value of SD (SD_diff_) was not reported, it was obtained 1) by multiplying SEs of means by the square root of the sample size when standard errors (SEs) of the means were reported, or 2) with SDs at the baseline (SD_baseline_) and post-intervention SD (SD_post_) in addition to the within-groups bivariate correlation coefficient (r) ^26^:

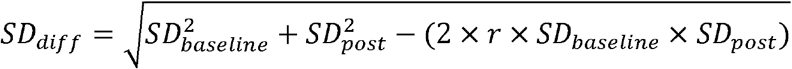

The I² statistic was employed for evaluating heterogeneity and a standard Chi^2^ test was employed for detecting whether significant heterogeneity existed. Heterogeneity was statistically significant at P < 0.10 after due consideration of I² statistic, of which a value greater than 50% was considered substantial heterogeneity ^27^. The random-effects model was applied where the evidence of heterogeneity was found.

The comparison was established between PEMFs and placebo control or exercise in the meta-analysis. The subgroup analysis was conducted to detect the effectiveness relative to different follow-up timepoints (postintervention; follow-up at 12, 24 weeks from baseline). To evaluate the quality and consistency of pooled results, the sensitivity analysis was conducted by deleting each included study. Where the data allowed, assessment of publication bias was performed. All tests were two-tailed, and P < 0.05 was considered statistically significant.

## RESULTS

### Flow of studies through the review

In total, 806 articles were identified by our search, of which 124 duplicate articles were removed. Based on title and abstract screening, 632 of these articles were excluded. Full texts of 50 articles were read, a further 42 articles were excluded, remaining 8 articles included in the data extraction and analysis of the review (Figure 1) ^28-35^.

**Figure 1.**
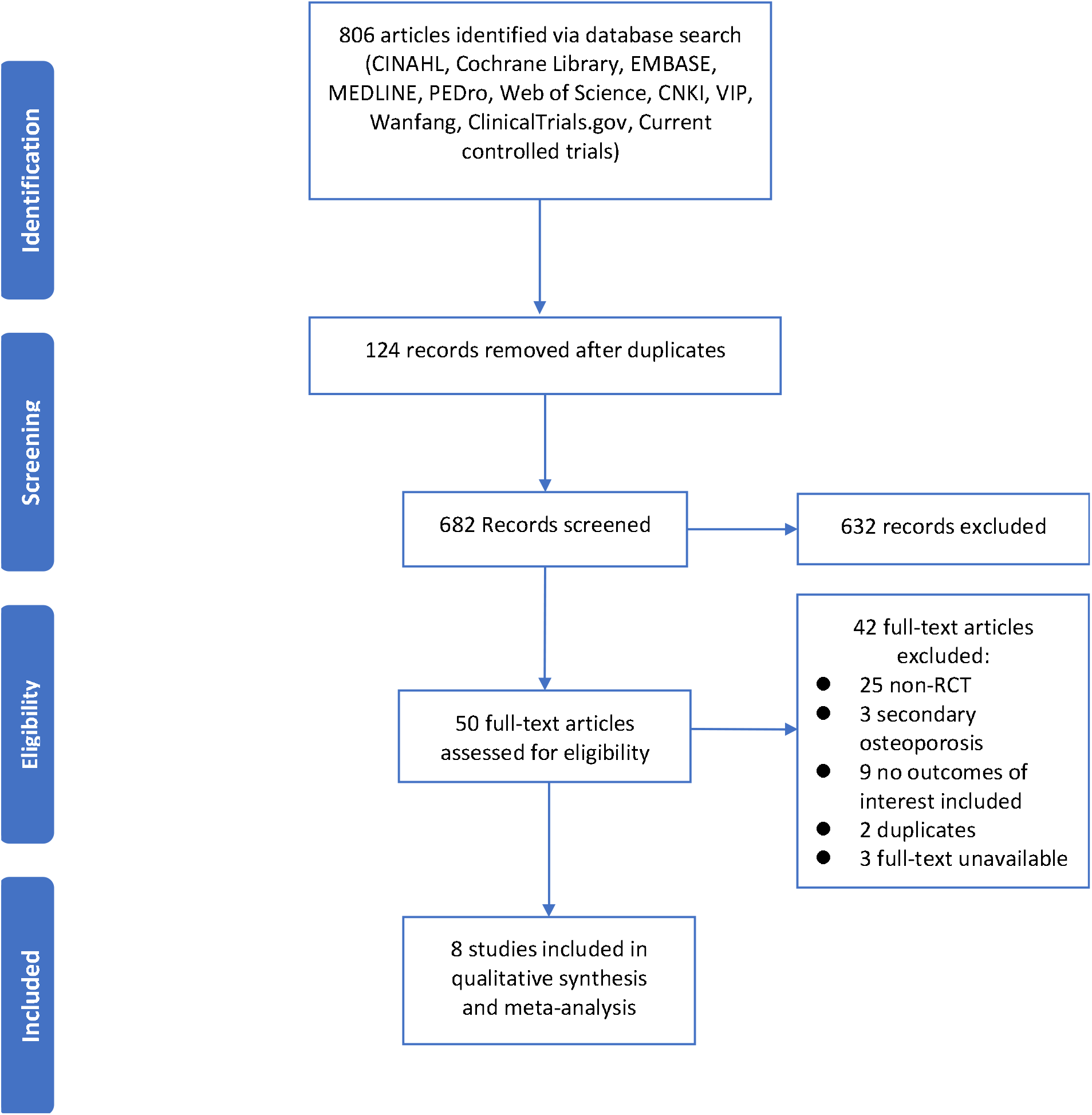
PRISMA flow of studies through the review.

### Characteristics of studies

#### Quality

All included studies achieved PEDro scores over 5, among which five studies achieved the score over 8 (Table 1)^28,29,31-33^. Of the 8 included studies, 2 studies were rated as ‘low risk of bias’ in all domains^31,32^, and other studies were classified as ‘unclear risk of bias’ for at least 1 aspect or ‘high risk of bias’ for at least 2 aspects. In results of GRADE, the quality of the evidence for the comparison between PEMFs versus placebo control was low or moderate, and that for the comparison between PEMFs versus exercise was very low. The results of the risk of bias and GRADE are presented in the Supplementary Tables and Figures.

**Table 1.**
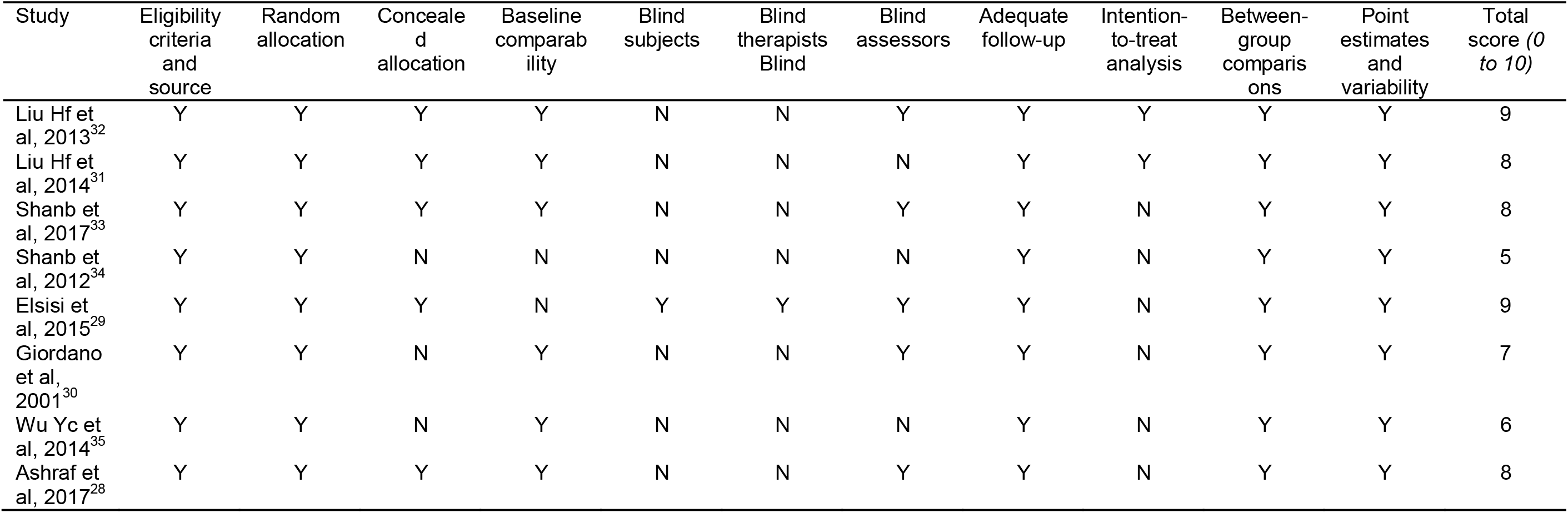
PEDro scores of included studies.

| Study | Eligibility<br>criteria<br>and<br>source | Random<br>allocation | Concealed<br>allocation | Baseline<br>comparability | Blind<br>subjects | Blind<br>therapists<br>Blind | Blind<br>assessors | Adequate<br>follow-up | Intention-to-treat<br>analysis | Between-<br>group<br>comparisons | Point<br>estimates<br>and<br>variability | Total<br>score (0<br>to 10) |
| --- | --- | --- | --- | --- | --- | --- | --- | --- | --- | --- | --- | --- |
| Liu Hf et al, 2013<sup>32</sup> | Y | Y | Y | Y | N | N | Y | Y | Y | Y | Y | 9 |
| Liu Hf et al, 2014<sup>31</sup> | Y | Y | Y | Y | N | N | N | Y | Y | Y | Y | 8 |
| Shanb et al, 2017<sup>33</sup> | Y | Y | Y | Y | N | N | Y | Y | N | Y | Y | 8 |
| Shanb et al, 2012<sup>34</sup> | Y | Y | N | N | N | N | N | Y | N | Y | Y | 5 |
| Elsisi et al, 2015<sup>29</sup> | Y | Y | Y | N | Y | Y | Y | Y | N | Y | Y | 9 |
| Giordano et al, 2001<sup>30</sup> | Y | Y | N | Y | N | N | Y | Y | N | Y | Y | 7 |
| Wu Yc et al, 2014<sup>35</sup> | Y | Y | N | Y | N | N | N | Y | N | Y | Y | 6 |
| Ashraf et al, 2017<sup>28</sup> | Y | Y | Y | Y | N | N | Y | Y | N | Y | Y | 8 |

#### Participants

In total, data were extracted for 396 participants, comprising 183 participants in PEMFs group and 213 participants in placebo control (alendronate/pharmacological therapy) or exercise group. Mean age ranged from 56.3 to 70 years, with a gender ratio of 49 to 347. Participant characteristics are detailed in Table 2.

**Table 2.** Characteristics of the included studies.

| Study | Participant |  |  |  | Intervention |  | Outcome measures |  |  | Follow-up |
| --- | --- | --- | --- | --- | --- | --- | --- | --- | --- | --- |
|  | Sample size | Study arms | Age (y) | Male/Female | Type of intervention | Dose per protocol<sup>a</sup> | Primary | Secondary | Efficacy/Effect |  |
| Liu Hf et al, 2013<sup>32</sup> | 41 | 1. PEMFs (n=20)<br>2. AL (n=21) | 61.70 (1.34)<br>63.14 (4.32) | 0/41 | PEMFs | 3.82 mT, 8 Hz, 40min/session for a total of 30 sessions. | BMDL | BMDF; 25(OH)D; LE MMT; BBS | Effective as alendronate. | 24 weeks |
| Liu Hf et al, 2014<sup>31</sup> | 84 | 1. PEMFs (n=44)<br>2. AL (n=40) | 62.41 (5.47)<br>62.63 (4.54) | 0/84 | PEMFs | 3.8 mT, 8-12 Hz, 40min/session for a total of 30 sessions. | BMDL; BMDF | VAS; TUG; BBS | Effective as alendronate. | 72 weeks |
| Shanb et al, 2017<sup>33</sup> | 68 | 1. PEMFs (n=25)<br>2. WBV (n=25)<br>3. AL (n=18) | 63.9 (3.9)<br>64.1 (4.4)<br>64.5 (4.03) | 23/45 | PEMFs | 5 mT, 33 Hz, 50 min/session, 2 sessions/week for 16 weeks. | BMDL; BMDF | Serum calcium and vitamin D | Effective in conjunction with pharmacological treatment. | N/A |
| Shanb et al, 2012<sup>34</sup> | 30 | 1. PEMFs (n=15)<br>2. Exercise (n=15) | 60-70 | 0/30 | PEMFs | 5 mT, 33 Hz, 50 min/session, 3 sessions/week for 12 weeks. | BMDL; BMDF | N/A | Effective as exercise. | N/A |
| Elsisi et al, 2015<sup>29</sup> | 30 | 1. PEMFs (n=15)<br>2. CRT-Exercise (n=15) | 64.73 (3.08)<br>65.13 (2.44) | 0/30 | PEMFs | 5 mT, 33 Hz, 30 min/session, 3 sessions/week for 12 weeks. | BMDL; BMDF | N/A | More effective than CRT exercise. | N/A |
| Giordano et al, 2001<sup>30</sup> | 40 | 1. PEMFs (n=20)<br>2. Placebo (n=20) | 56.3 (4.0)<br>55.9 (3.1) | 0/40 | PEMFs | 1.2 mT, 100 Hz, 60 min/session, 3 sessions/week for 12 weeks. | BMDL; BMDF | Serum and urinary calcium, phosphate; Serum ALP, osteocalcin and PICP; Urinary hydroxyproline | Not effective in increasing BMD, but effective in stimulating osteogenesis. | 1 month |
| Wu Yc et al, 2014<sup>35</sup> | 43 | 1. PEMFs (n=24)<br>2. AL (n=19) | 59.08 (4.65)<br>59.53 (5.40) | 0/43 | PEMFs | 3.8 mT, 8-12 Hz, 40min/session for a total of 30 sessions. | TUG; BBS; Balance function | N/A | Effective as alendronate. | N/A |
| Ashraf et al, 2017<sup>28</sup> | 60 | 1. PEMFs (n=20)<br>2. LLLT (n=20)<br>3. Med (n=20) | 59.85 (2.35)<br>60.20 (2.17)<br>60.00 (2.62) | 26/34 | PEMFs | 4 mT, 33 Hz, 30 min/session, 3 sessions/week for 12 weeks. | BMDL | N/A | More effective than LLLT and AL group. | N/A |

<sup>a</sup>Waveforms; Intensity; Frequency; Duration (weeks).
<sup>b</sup>Abbreviations. PEMFs=Pulsed electromagnetic fields, AL=alendronate, WBV=Whole body vibration, CRT=Circuit weight training, LLLT=low-level laser therapy, mT=millitesla, Hz=Hertz, BMDL=lumbar bone mineral density, BMDF=femur bone mineral density, 25(OH)D=serum 25OH vitamin, LE MMT=total lower-extremity manual muscle test, BBS=Berg Balance Scale, VAS=Visual Analogue Scale, N/A=not applicable, TUG=Timed Up and Go, ALP=Alkaline Phosphatase, PICP=Procollagen type I carboxy-terminal propeptide, Med=Medication.

#### Intervention

The frequency and intensity of PEMFs exposure varied at 8-100 Hz and 1.2-5 mT separately. A range of 30-36 PEMFs sessions, with 30-60 min/session, were prescribed for participants, and 4 to 72 weeks follow-up were conducted across all studies. Control intervention types included first-line pharmacological agents (e.g., Alendronate, intake of Vitamin D and Calcium) and exercise (e.g., whole body vibration and resistance training). The intervention characteristics of the included studies are detailed in the Table 2.

#### Outcome measures

The outcome measures in each study with the categories of bone mass and functional assessments are detailed in Table 2.

### Effect of intervention I: PEMFs versus control group

#### Bone Mineral Density (BMD)

Five studies (study population, N=248) ^28,30-33^and three studies (study population, N=124) ^30,32,33^reported data on percentage change in BMD at the lumbar and femur neck respectively after intervention directly (Figure 2). Low certainty evidence showed that PEMFs has no effect on BMD at the lumbar (MD 0.01; CI -0.04 to 0.06) and femur neck (MD 0.01; CI -0.02 to 0.04) with no statistically significant heterogeneity (lumbar: I²= 0%, P =0.48; femur neck: I² = 0%, P = 0.79). Two studies (study population, N=125) ^31,32^performed follow-ups at 12 and 24 weeks from baseline on percentage change in BMD at the lumbar (Figure 2). Moderate certainty evidence showed that there is no statistically significant effect on BMD at the lumbar at the 12 weeks follow-up (MD -0.01; CI -0.28 to 0.25) and the 24 weeks follow-up (MD -0.02; CI -0.30 to 0.26) with no statistically significant heterogeneity (12 weeks: I² = 0%, P = 1.00; 24 weeks: I² = 0%, P = 0.98). No significant changes in heterogeneity and overall effect were observed in the sensitivity analysis.

**Figure 2.**
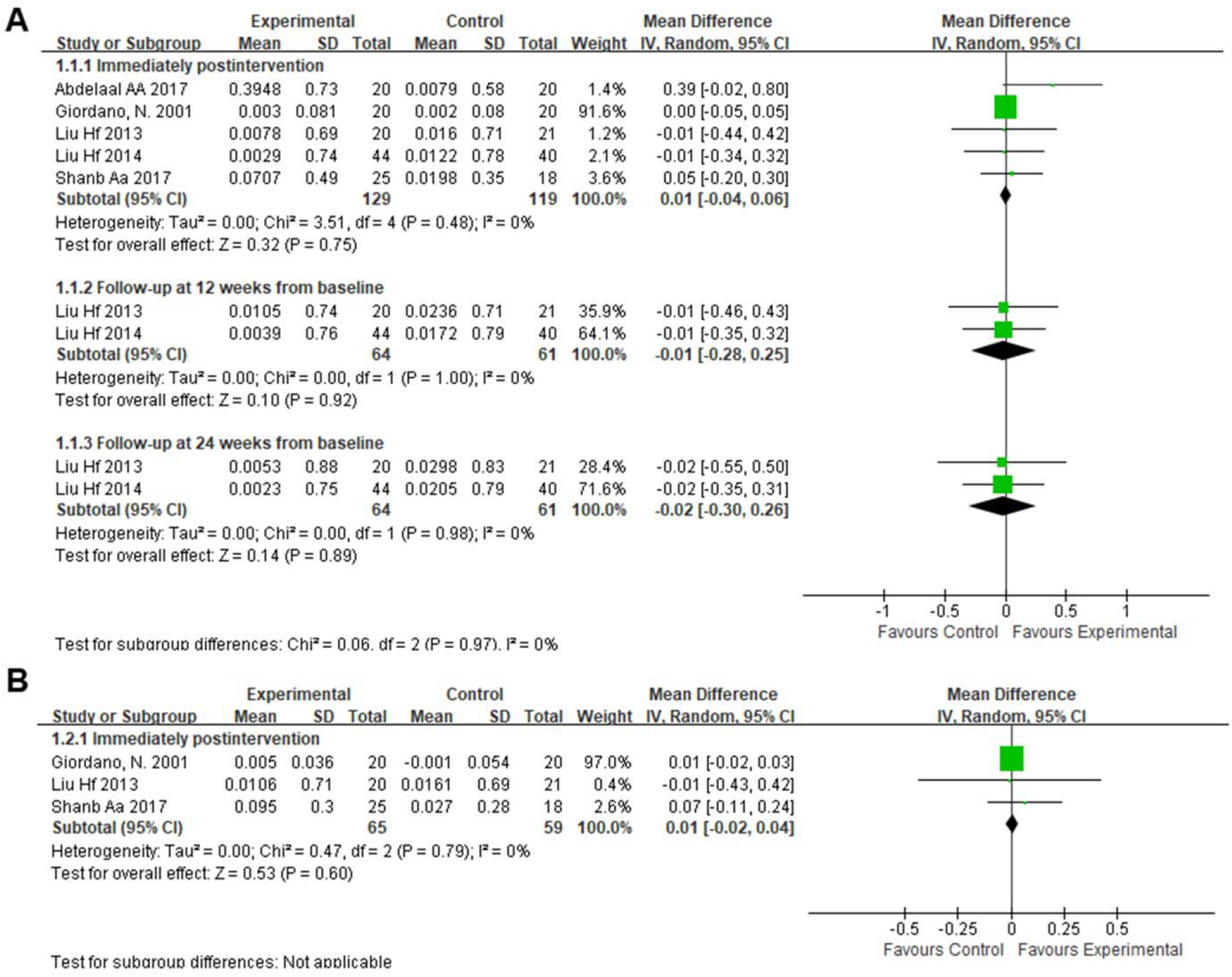
Forest plot analysis of the effects of PEMFs on BMD at the lumbar (A) and femur neck (B) compared with placebo control. Data are presented as mean difference (MD) between treatment and control groups with a 95% confidence interval (CI). SD = Standard Deviation; BMD = Bone Mineral Density; PEMFs = Pulsed Electromagnetic Fields.

#### Berg Balance Scale (BBS)

Three studies (study population, N=168) ^31,32,35^conducted the assessment of BBS after intervention directly (Figure 3). Low certainty evidence showed that there is no statistically significant effect on percentage change in BBS (MD 0.01; CI -0.09 to 0.11) with no statistically significant heterogeneity (I² = 0%, P = 1.00). Two studies (study population, N=125) ^31,32^carried out follow-ups at 12 and 24 weeks from baseline on percentage change in BBS (Figure 3). Moderate certainty evidence showed that no statistically significant effect is detected on BBS at the 12 weeks (MD 0.00; CI -0.13 to 0.14) and 24 weeks follow-up (MD -0.00; CI -0.15 to 0.14) with evidence of no statistically significant heterogeneity (12 weeks: I² = 0%, P = 0.99; 24 weeks: I² = 0%, P = 0.99). No significant changes in heterogeneity and overall effect were observed in the sensitivity analysis.

**Figure 3.**
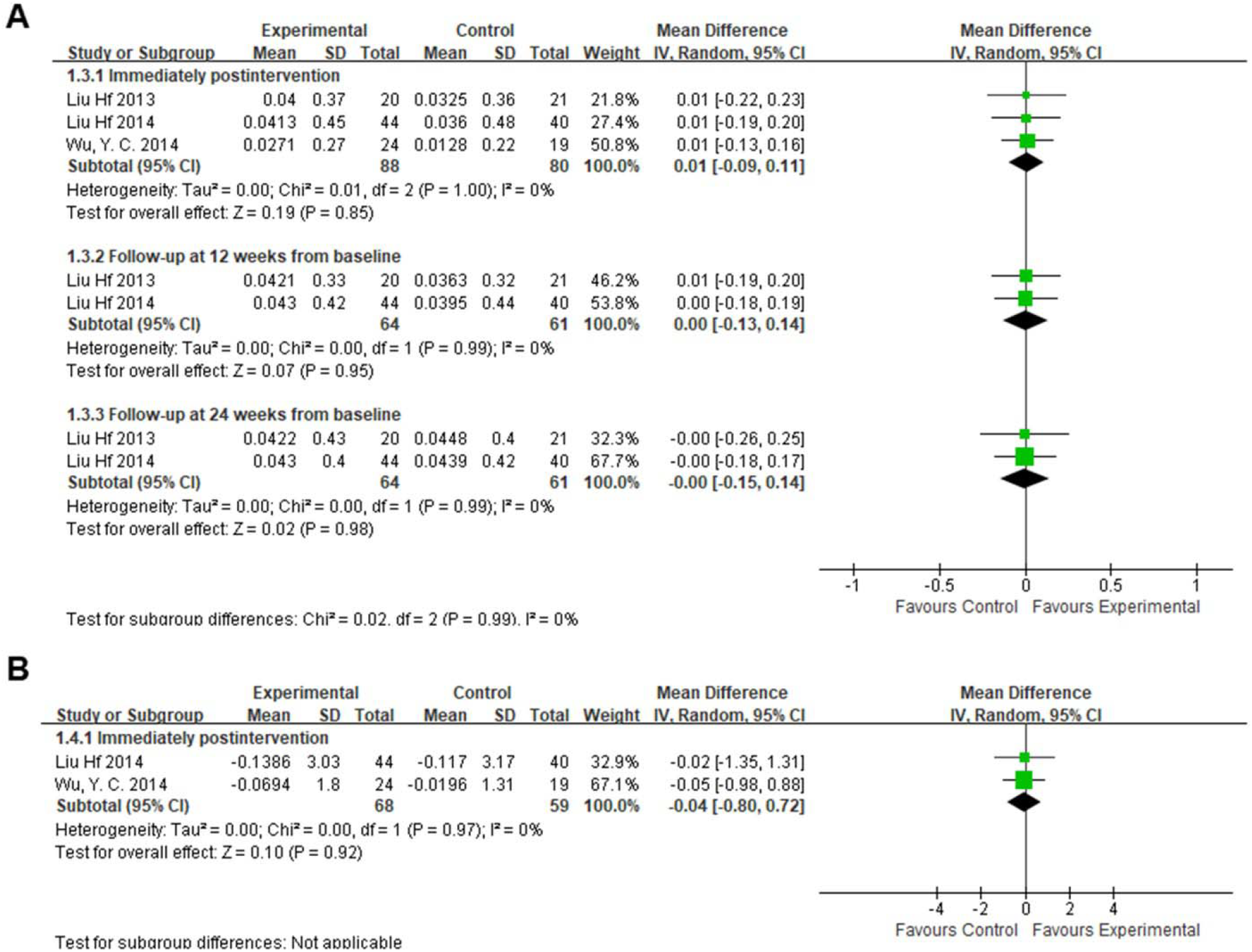
Forest plot analysis of the effects of PEMFs on balance function measured by BBS (A) and TUG test (B) compared with placebo control. Data are presented as mean difference (MD) between treatment and control groups with a 95% confidence interval (CI). SD = Standard Deviation; BBS = Berg Balance Scale; TUG = Timed Up and Go; PEMFs = Pulsed Electromagnetic Fields.

#### Timed Up and Go (TUG) test

Two studies (study population, N=127) ^31,35^assessed the percentage change in TUG test after intervention directly (Figure 3). Low certainty evidence showed that there is no statistically significant effect on TUG (MD -0.04; CI -0.80 to 0.72) with no statistically significant heterogeneity (I² = 0%, P = 0.97). No significant changes in heterogeneity and overall effect were observed in the sensitivity analysis.

### Effect of intervention II: PEMFs versus exercise group

Three studies (study population, N=110) ^29,33,34^investigated the effect of PEMFs on percentage change in BMD at the lumbar and femur neck respectively after intervention directly (Figure 4). Very low certainty evidence showed that PEMFs has small but significant effect on BMD at the femur neck (MD 0.10; CI 0.01 to 0.20), and no effect on BMD at the lumbar (MD 0.15; CI -0.04 to 0.35), both with no statistically significant heterogeneity respectively (femur neck: I² = 28%, P = 0.25; I² = 47%, P = 0.15). In the sensitivity analysis, one study ^29^was found to be a contributor to results of non-significant heterogeneity and the effect on BMD at the femur neck. After excluding the study, the heterogeneity was reduced to 0% (femur neck: P = 0.96; P = 0.86), and the small but significant effect on BMD at the femur neck was eliminated (MD 0.00; CI -0.15 to 0.16).

**Figure 4.**
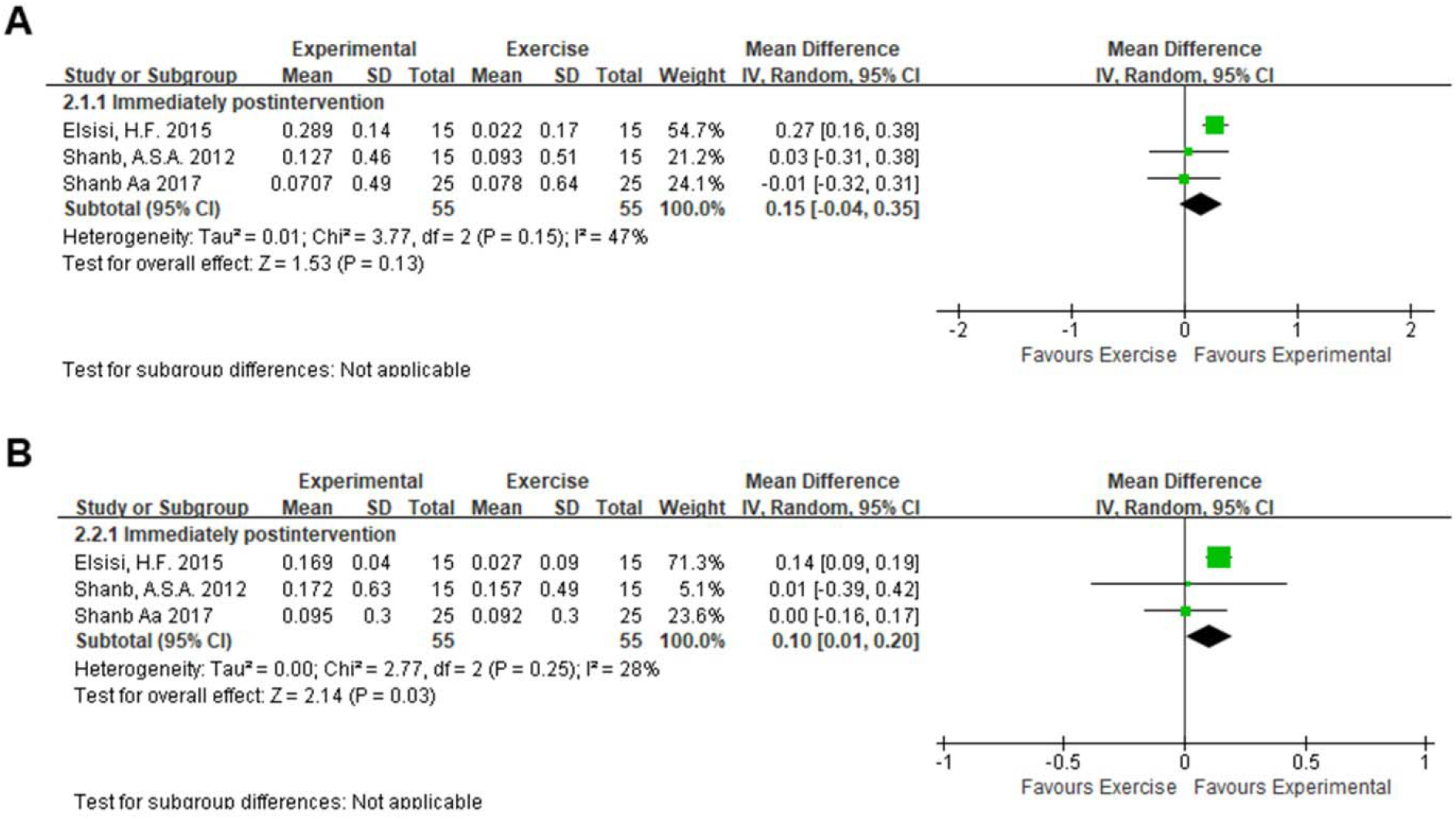
Forest plot analysis of the effects of PEMFs on BMD at the lumbar (A) and femur neck (B) compared with exercise group. Data are presented as mean difference (MD) between treatment and control groups with a 95% confidence interval (CI). SD = Standard Deviation; BMD = Bone Mineral Density; PEMFs = Pulsed Electromagnetic Fields.

## DISCUSSION

This systematic review and meta-analysis of 8 studies involving 396 participants demonstrated PEMFs as a physical therapy was non-inferior to conventional pharmacological agents in preventing the decline of BMD and balance function for the management of primary osteoporosis in older adults. Meanwhile, we also found that PEMFs might be slightly more effective in increasing BMD than exercise. According to our knowledge, no systematic review and meta-analysis was initiated before, except one network meta-analysis exploring effects of nonpharmacological interventions including PEMFs on balance function only ^36^, and several narrative reviews including clinical studies were retrieved ^11,37,38^. Our results are in consistent with findings from previous reviews^11,36,37^ that PEMFs achieved positive effects on BMD and balance function for older adults with primary osteoporosis, implicating that PEMFs may potentially become a promising treatment option.

Bisphosphonates and exercise were both identified as the first-line interventions for the management of osteoporosis in the latest evidence-based guideline ^10^. Our study established comparisons between PEMFs and active placebo or exercise based on groups set by included studies, and sub-group analysis was stratified by different intervals between the baseline, post-intervention, and follow-ups. BMD, as a surrogate measure for therapeutic effectiveness, can be assessed by various methods, among which dual x-ray absorptiometry (DXA) was proved to be reliable in the diagnosis of osteoporosis and have relatively good responsiveness in RCT ^39,40^. Our study demonstrated that there was no difference between PEMFs and active placebo in improving BMD at the lumbar and femur neck in all sub-group meta-analysis, which suggests PEMFs is nearly effective as pharmacological agents for osteoporosis, and the effect could last for at least 24 weeks. Moreover, the effects of PEMFs versus exercise on BMD at the lumbar was not significant, while that was detected as small but significant on BMD at the femur neck. Our results were considered fairly stable as the sensitivity analysis only detected one study ^29^affecting the heterogeneity and effect of PEMFs versus exercise on BMD at the femur neck. This is likely due to variations in exercise characteristics (e.g., duration, training load and training volume). Therefore, this part of results should be interpreted with caution.

Impaired balance function is an important risk factor increasing the incidence of falling and fracture, which is modifiable by balance-improving interventions ^41^. In line with results on BMD, no statistically significant difference was observed for balance function measured by BBS and TUG after the intervention of PEMFs versus active placebo. In the sub-group analysis, the effect of PEMFs on BBS at 12, 24 weeks follow-up was also not statistically significant, confirming that PEMFs is as effective as conventional pharmacological agents in improving balance function which could last for 24 weeks. In consistent with a previous systematic review ^36^, it reported that PEMFs exert positive effect on BBS and TUG tests reflecting balance function. However, the network meta-analysis was conducted to further compare the effects of five interventions on balance function with the conclusion that balance and strength training was better than other interventions. Only one study using data on PEMFs was included in the analysis, compared to that we included 3 studies, and no study was ever conducted to directly compare the effect of PEMFs versus other non-pharmacological interventions on balance function, combining these two may explain, in part, the conflicting results. Furthermore, our sensitivity analysis detecting no changes in levels of heterogeneity and effect confirmed our results as relatively robust.

To overcome the shortcomings of the “statistically significant difference”, the minimum clinically important difference (MCID) defined as “the smallest change that is important to patients” is employed to generate a threshold value for such change ^42^. Any patient whose responses help them reach the MCID threshold is considered as responders. Thus, a certain proportion of responders to the total participants involved in a trialed intervention indicates the likelihood of patients under the same condition also responding favorably to the same intervention ^43^. However, no definite consensus reached on the MCID of BMD and balance function. Some evidence showed that changes by 2-5% at the lumbar and 8% at the proximal femur ^44^, a point-drop in BBS associated with a 3-4% increase in risk of falling ^45^, and an improvement of 2-3 seconds in TUG test were considered as MCID for the older population ^46^. In our study, no study included used the MCID and responder rate to evaluate the effect of PEMFs, thus, it is hard to determine the clinical importance of improvements achieved by PEMFs on BMD and balance function compared with placebo and exercise. Furthermore, the successful treatment of osteoporosis is prevention of fractures, while no treatment can completely eliminate fracture risk ^10^. Although a certain increase in BMD and the improvement in balance function for osteoporosis may result in a reduction in fracture risk, no risk and number of incident fracture were reported in studies included in our review. Therefore, studies with reporting measures of MCID, fracture risk and incidence in the future are required to further confirm that the effect of PEMFs in treating osteoporosis is of clinical importance to clinicians and patients.

This review has several strengths. To date, we are the first to conduct a systematic review and meta-analysis to explore the effect of PEMFs on BMD and balance function for primary osteoporosis. Secondary, the results of our study were fairly stable due to studies included were of appropriate methodological quality, and no significant heterogeneity was found in the analysis. Furthermore, a key finding of this review was that PEMFs as an intervention alone achieved an effect non-inferior to first-line treatment (pharmacological agents and exercise) on improvements in BMD and balance function for older adults with primary osteoporosis. At last, a librarian familiar with the development of searches and a mechanism of living systematic review were involved over a course of 3 years to ensure no study was missed in compliance with the study protocol.

There are several limitations to this review that deserve consideration. The primary limitation of our review was the limited number of included studies with only 8 studies comprising of 396 participants for the analysis, from which the level of evidence generated were moderate to very low. In some sub-group analyses, only 2-3 studies were included from which some uncertainty in the results interpretation may exist based on data extracted. In addition, as the maximum PEMF treatment session lasted for 16 weeks and the longest follow-up was 24 weeks for the analysis, the error among the minimum percentage change in BMD and balance function may stay undetectable and further deepened the uncertainty ^44^. Furthermore, the clinical relevance of the findings was limited, combined with no study reported data on MCID, responder rate and incident fracture and only two studies ^31,32^included used intention-to-treat (ITT) analysis. According to accumulated evidence ^11,47^, the parameters of non-pharmacological interventions, including intensity, frequency, and duration, were critical to impact changes in outcome measures for osteoporosis, while our study could not conduct the sub-group analysis based on different parameters due to limited information retrieved. Another potential limitation was the systematic search was limited to English and Chinese manuscripts available in full text, and some relevant trials may be missed.

## CONCLUSION

In summary, moderate to very low certainty evidence showed that PEMFs as an intervention alone has positive effects non-inferior to first-line treatment (pharmacological agents and exercise) on BMD and balance function in older adults with primary osteoporosis and should be considered as a promising option in the management of osteoporosis. Although uncertainty about responses to the intervention and changes in outcome measures may be existed but undetectable, our findings may still be fairly stable as we consistently found similar results in the primary and sensitivity analyses. For further deeply confirming the effect of PEMFs for osteoporosis, adding endpoints like fracture risk and incidence, and outcome measures like MCID and responder rate to the core collection are necessary. In the future, researchers planning a PEMFs study should optimize the study design, with taking factors not limited to undetectable errors under outcomes, parameters of interventions, longer follow-up period, larger sample size and ITT analysis into consideration, to generate high certainty evidence.

## Supporting information

The search strategies, Table S (1, 2), Figure S (1, 2), and PRISMA checklist can be found in the file.

## Data Availability

No additional data available.

## DECLARATIONS

## Acknowledgements

We are grateful for the partnership and support from the library of Sichuan University for the development of search strategy, also and the clinical research and statistics office of West China Hospital for data processing and analysis.

## Availability of Data and Materials

Not applicable

## Authors’ contributions

All authors have made substantial contributions to the study design. SYZ and YL have contributed to the ongoing data collection. All listed authors have contributed to the drafting of the manuscript.

## Box of inclusion criteria for eligible studies

### Box 1.

Inclusion criteria

#### Design

- Randomised or quasi-randomised controlled trial
- Published in a peer-reviewed journal
- Full text available in Chinese and English

#### Participants

- Healthy older adults (including those with previous fractures) aged over 50 years with primary osteoporosis

#### Intervention

- Electromagnetic fields with pulsed signal and extremely low frequencies (between 5 and 300 Hz)

#### Outcome measures

- Primary outcomes: bone mass, number of incident fractures
- Secondary outcomes: Functional assessments, quality of life, adverse events

#### Comparisons

- PEMFs versus sham/nothing
- PEMFs versus placebo/pharmacological agents
- PEMFs versus exercise/other physical agent or intervention
- PEMFs plus other intervention versus other intervention

