## Supplementary material for "Pulsed electromagnetic fields may be effective for the management of primary osteoporosis: a systematic review and meta-analysis": The search strategies, Table S (1, 2), Figure S (1, 2), and PRISMA checklist can be found in the file.

**Appendix I. Search strategy**

**Databases: MEDLINE**

1. OSTEOPOROSIS/

2. osteoporo$.mp.

3. osteopenia.mp.

4. bone density/

5. bone densit$.mp.

6. bone loss$.mp.

7. bone mass$.mp.

8. bone mineral densit$.mp.

9. bone mineral content$.mp.

10. bone defect$.mp.

11. bone deminerali?ation.mp.

12. bone mineral$.mp.

13. bone strength.mp.

14. decalcifi$.mp.

15. deminerali?ed bone.mp.

16. bone age.mp.

17. 1 or 2 or 3 or 4 or 5 or 6 or 7 or 8 or 9 or 10 or 11 or 12 or 13 or 14 or 15 or 16

18. Electromagnetic Field$.mp.

19. exp Electromagnetic Fields/

20. exp Magnetic Fields/

21. Magnetic Field$.mp.

22. exp Magnetic Field Therapy/

23. exp Electric Stimulation Therapy/

24. Magnetic Field$ Therapy.mp.

25. PEMF$.mp.

26. 18 or 19 or 20 or 21 or 22 or 23 or 24 or 25

27. ((randomized controlled trial or controlled clinical trial).pt. or randomized.ab. or randomised.ab. or placebo.ab. or drug therapy.fs. or randomly.ab. or trial.ab. or groups.ab.) not (exp animals/ not humans.sh.).af.

28. 17 and 26 and 27

29. randomized controlled trial.pt.

30. controlled clinical trial.pt.

31. randomi?ed.ab.

32. placebo.ab.

33. clinical trials as topic.sh.

34. randomly.ab.

35. trial.ti.

36. 29 or 30 or 31 or 32 or 33 or 34 or 35

37. 17 and 26 and 36

**Databases: EMBASE**

1. osteoporosis/

2. osteoporo$.mp.

3. osteopenia.mp.

4. bone density/

5. bone mass/

6. bone densit$.mp.

7. exp bone/

8. bone loss$.mp.

9. bone mass$.mp.

10. bone mineral/

11. bone mineral densit$.mp.

12. bone mineral content$.mp.

13. 1 or 2 or 3 or 4 or 5 or 6 or 7 or 8 or 9 or 10 or 11 or 12

14. Electromagnetic Field$.mp.

15. exp pulsed electric field/

16. pulsed electric field$.mp.

17. exp magnetotherapy/

18. Randomized Controlled Trial/

19. Double Blind Procedure/

20. Single Blind Procedure/

21. Triple Blind Procedure/

22. randomi?ed.ti,ab.

23. randomisation/

24. Placebo/

25. placebo$.mp.

26. ((controlled or comparative or placebo or randomi?ed) adj3 (trial or study)).mp.

27. (random$ adj7 (allocat$ or allot$ or assign$ or basis$ or divid$ or order$)).mp.

28. ((singl$ or doubl$ or trebl$ or tripl$) adj7 (blind$ or mask$)).mp.

29. 18 or 19 or 20 or 21 or 22 or 23 or 24 or 25 or 26 or 27 or 28

30. exp electromagnetic field/

31. electromagnetism/

32. exp low frequency electrotherapy/

33. electrotherapy/

34. 14 or 15 or 16 or 17 or 30 or 31 or 32 or 33

35. 13 and 29 and 34

**Databases: CCTR**

1. Osteoporosis/

2. osteoporo$.mp.

3. osteopenia.mp.

4. bone density/

5. bone densit$.mp.

6. exp "Bone and Bones"/

7. bone loss$.mp.

8. bone mass$.mp.

9. bone mineral content$.mp.

10. bone mineral densit$.mp.

11. bone strength.mp.

12. bone mineral$.mp.

13. 1 or 2 or 3 or 4 or 5 or 6 or 7 or 8 or 9 or 10 or 11 or 12

14. randomized controlled trial.pt.

15. controlled clinical trial.pt.

16. placebo.ab.

17. trial.ti.

18. randomly.ab.

19. clinical trials as topic.sh.

20. randomi?ed.ab.

21. 14 or 15 or 16 or 17 or 18 or 19 or 20

22. exp Electromagnetic Fields/

23. exp Magnetic Field Therapy/

24. exp Magnetic Fields/

25. exp Pulsed Radiofrequency Treatment/

26. Electromagnetic Field$.mp.

27. Magnetic Field$.mp.

28. Magnetic Field$ Therapy.mp.

29. 22 or 23 or 24 or 25 or 26 or 27 or 28

30. 13 and 21 and 29

**Databases: PEDro**

Osteoporo* or bone in Abstract & Title AND electrotherapies, heat, cold in Therapy

**Databases: Pubmed/CNKI (Chinese)/WANFANG (Chinese)**

(((((((("Electromagnetic Fields"[Mesh]) OR Electromagnetic Field[Title/Abstract]) OR Field, Electromagnetic[Title/Abstract]) OR Fields, Electromagnetic[Title/Abstract]) OR pulsed electromagnetic fields[Title/Abstract]) OR PEMF[Title/Abstract])) AND (((("Osteoporosis"[Mesh]) OR bone loss[Title/Abstract]) OR OP[Title/Abstract]) OR Osteoporoses[Title/Abstract]))

**Appendix II. Tables**

**Table S1**

Summary of findings for comparison between PEMFs and placebo control

| **Electromagnetic fields compared to Control for preventing and treating osteoporosis: a systematic review and meta-analysis** | | | | | | |
| --- | --- | --- | --- | --- | --- | --- |
| **Patient or population:** patients with preventing and treating osteoporosis: a systematic review and meta-analysis **Settings:**  **Intervention:** Electromagnetic fields **Comparison:** Control | | | | | | |
| **Outcomes** | **Illustrative comparative risks* (95% CI)** | | **Relative effect (95% CI)** | **No of Participants (studies)** | **Quality of the evidence (GRADE)** | **Comments** |
|  | Assumed risk | Corresponding risk |  |  |  |  |
|  | **Control** | **Electromagnetic fields** |  |  |  |  |
| **Bone Mineral Density % Change: Lumbar - Immediately postintervention** Dual-energy X-ray absorptiometry |  | The mean bone mineral density % change: lumbar - immediately postintervention in the intervention groups was **0.01 higher** (0.04 lower to 0.06 higher) |  | 248 (5 studies) | ⊕⊕⊝⊝ **low**^1^ |  |
| **Bone Mineral Density % Change: Lumbar - Follow-up at 12 weeks from baseline** Dual-energy X-ray absorptiometry |  | The mean bone mineral density % change: lumbar - follow-up at 12 weeks from baseline in the intervention groups was **0.01 lower** (0.28 lower to 0.25 higher) |  | 125 (2 studies) | ⊕⊕⊕⊝ **moderate**^2^ |  |
| **Bone Mineral Density % Change: Lumbar - Follow-up at 24 weeks from baseline** Dual-energy X-ray absorptiometry |  | The mean bone mineral density % change: lumbar - follow-up at 24 weeks from baseline in the intervention groups was **0.02 lower** (0.3 lower to 0.26 higher) |  | 125 (2 studies) | ⊕⊕⊕⊝ **moderate**^2^ |  |
| **Bone Mineral Density % Change: Femur neck - Immediately postintervention** Dual-energy X-ray absorptiometry |  | The mean bone mineral density % change: femur neck - immediately postintervention in the intervention groups was **0.01 higher** (0.02 lower to 0.04 higher) |  | 124 (3 studies) | ⊕⊕⊝⊝ **low**^3^ |  |
| **BBS - Immediately postintervention** The Berg Balance Scale |  | The mean bbs - immediately postintervention in the intervention groups was **0.01 higher** (0.09 lower to 0.11 higher) |  | 168 (3 studies) | ⊕⊕⊝⊝ **low**^3^ |  |
| **BBS - Follow-up at 12 weeks from baseline** The Berg Balance Scale |  | The mean bbs - follow-up at 12 weeks from baseline in the intervention groups was **0 higher** (0.13 lower to 0.14 higher) |  | 125 (2 studies) | ⊕⊕⊕⊝ **moderate**^2^ |  |
| **BBS - Follow-up at 24 weeks from baseline** The Berg Balance Scale |  | The mean bbs - follow-up at 24 weeks from baseline in the intervention groups was **0 higher** (0.15 lower to 0.14 higher) |  | 125 (2 studies) | ⊕⊕⊕⊝ **moderate**^2^ |  |
| **TUG - Immediately postintervention** Timed Up and Go Test |  | The mean tug - immediately postintervention in the intervention groups was **0.04 lower** (0.8 lower to 0.72 higher) |  | 127 (2 studies) | ⊕⊕⊝⊝ **low**^3^ |  |
| *The basis for the **assumed risk** (e.g. the median control group risk across studies) is provided in footnotes. The **corresponding risk** (and its 95% confidence interval) is based on the assumed risk in the comparison group and the **relative effect** of the intervention (and its 95% CI).  **CI:** Confidence interval; | | | | | | |
| GRADE Working Group grades of evidence **High quality:** Further research is very unlikely to change our confidence in the estimate of effect.  **Moderate quality:** Further research is likely to have an important impact on our confidence in the estimate of effect and may change the estimate. **Low quality:** Further research is very likely to have an important impact on our confidence in the estimate of effect and is likely to change the estimate. **Very low quality:** We are very uncertain about the estimate. | | | | | | |
| ^1^ Downgraded for small samle size; unclear risk for allocation concealment, blinding of participants and personnel, blinding of outcome assessment, and selective reporting. ^2^ Downgraded for small samle size. ^3^ Downgraded for small samle size; unclear risk for allocation concealment, blinding of participants and personnel, and blinding of outcome assessment. | | | | | | |

**Table S2**

Summary of findings for comparison between PEMFs and exercise

| **Electromagnetic fields compared to Exercise for preventing and treating osteoporosis: a systematic review and meta-analysis** | | | | | | |
| --- | --- | --- | --- | --- | --- | --- |
| **Patient or population:** patients with preventing and treating osteoporosis: a systematic review and meta-analysis **Settings:**  **Intervention:** Electromagnetic fields **Comparison:** Exercise | | | | | | |
| **Outcomes** | **Illustrative comparative risks* (95% CI)** | | **Relative effect (95% CI)** | **No of Participants (studies)** | **Quality of the evidence (GRADE)** | **Comments** |
|  | Assumed risk | Corresponding risk |  |  |  |  |
|  | **Exercise** | **Electromagnetic fields** |  |  |  |  |
| **Bone Mineral Density % Change: Lumbar - Immediately postintervention** |  | The mean bone mineral density % change: lumbar - immediately postintervention in the intervention groups was **0.15 higher** (0.04 lower to 0.35 higher) |  | 110 (3 studies) | ⊕⊝⊝⊝ **very low**^1^ |  |
| **Bone Mineral Density % Change: Femur neck - Immediately postintervention** |  | The mean bone mineral density % change: femur neck - immediately postintervention in the intervention groups was **0.1 higher** (0.01 to 0.2 higher) |  | 110 (3 studies) | ⊕⊝⊝⊝ **very low**^1^ |  |
| *The basis for the **assumed risk** (e.g. the median control group risk across studies) is provided in footnotes. The **corresponding risk** (and its 95% confidence interval) is based on the assumed risk in the comparison group and the **relative effect** of the intervention (and its 95% CI).  **CI:** Confidence interval; | | | | | | |
| GRADE Working Group grades of evidence **High quality:** Further research is very unlikely to change our confidence in the estimate of effect.  **Moderate quality:** Further research is likely to have an important impact on our confidence in the estimate of effect and may change the estimate. **Low quality:** Further research is very likely to have an important impact on our confidence in the estimate of effect and is likely to change the estimate. **Very low quality:** We are very uncertain about the estimate. | | | | | | |
| ^1^ Downgraded for low-moderate heterogeneity; high risk for random sequence generation and allocation concealment; samll sample size; unclear risk for blinding of participants and personnel and blinding of outcome assessment. | | | | | | |

**Appendix III. Figures**

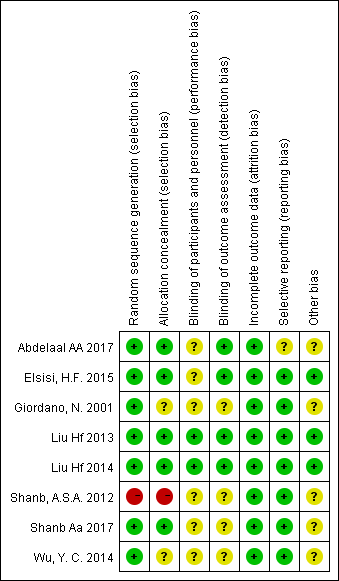

**Figure S1.** Cochrane Risk of bias summary: review authors' judgements about each risk of bias item for each included study.

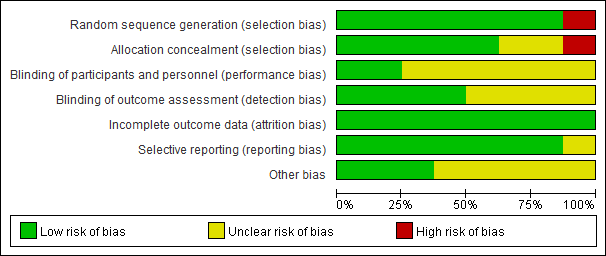

**Figure S2.** Cochrane Risk of bias summary: review authors' judgements about each risk of bias item presented as percentages across all included studies.

**Appendix IV. PRISMA checklist**

**PRISMA 2020 Main Checklist**

| **Topic** | **No.** | **Item** | **Location where item is reported** |
| --- | --- | --- | --- |
| **TITLE** |  |  |  |
| **Title** | 1 | Identify the report as a systematic review. | Page 1 |
| **ABSTRACT** |  |  |  |
| **Abstract** | 2 | See the PRISMA 2020 for Abstracts checklist |  |
| **INTRODUCTION** |  |  |  |
| **Rationale** | 3 | Describe the rationale for the review in the context of existing knowledge. | Page 4 |
| **Objectives** | 4 | Provide an explicit statement of the objective(s) or question(s) the review addresses. | Page 4-5 |
| **METHODS** |  |  |  |
| **Eligibility criteria** | 5 | Specify the inclusion and exclusion criteria for the review and how studies were grouped for the syntheses. | Page 5, 17 |
| **Information sources** | 6 | Specify all databases, registers, websites, organisations, reference lists and other sources searched or consulted to identify studies. Specify the date when each source was last searched or consulted. | Page 5 |
| **Search strategy** | 7 | Present the full search strategies for all databases, registers and websites, including any filters and limits used. | Page 5 and appendix files |
| **Selection process** | 8 | Specify the methods used to decide whether a study met the inclusion criteria of the review, including how many reviewers screened each record and each report retrieved, whether they worked independently, and if applicable, details of automation tools used in the process. | Page 5-6 |
| **Data collection process** | 9 | Specify the methods used to collect data from reports, including how many reviewers collected data from each report, whether they worked independently, any processes for obtaining or confirming data from study investigators, and if applicable, details of automation tools used in the process. | Page 5-7 |
| **Data items** | 10a | List and define all outcomes for which data were sought. Specify whether all results that were compatible with each outcome domain in each study were sought (e.g. for all measures, time points, analyses), and if not, the methods used to decide which results to collect. | Page 5-7 |
|  | 10b | List and define all other variables for which data were sought (e.g. participant and intervention characteristics, funding sources). Describe any assumptions made about any missing or unclear information. | Page 5-7 |
| **Study risk of bias assessment** | 11 | Specify the methods used to assess risk of bias in the included studies, including details of the tool(s) used, how many reviewers assessed each study and whether they worked independently, and if applicable, details of automation tools used in the process. | Page 6 |
| **Effect measures** | 12 | Specify for each outcome the effect measure(s) (e.g. risk ratio, mean difference) used in the synthesis or presentation of results. | Page 6-7 |
| **Synthesis methods** | 13a | Describe the processes used to decide which studies were eligible for each synthesis (e.g. tabulating the study intervention characteristics and comparing against the planned groups for each synthesis (item 5)). | Page 7-8 |
|  | 13b | Describe any methods required to prepare the data for presentation or synthesis, such as handling of missing summary statistics, or data conversions. | Page 7-8 |
|  | 13c | Describe any methods used to tabulate or visually display results of individual studies and syntheses. | Page 7-8 |
|  | 13d | Describe any methods used to synthesize results and provide a rationale for the choice(s). If meta-analysis was performed, describe the model(s), method(s) to identify the presence and extent of statistical heterogeneity, and software package(s) used. | Page 7-8 |
|  | 13e | Describe any methods used to explore possible causes of heterogeneity among study results (e.g. subgroup analysis, meta-regression). | Page 7-8 |
|  | 13f | Describe any sensitivity analyses conducted to assess robustness of the synthesized results. | Page 7-8 |
| **Reporting bias assessment** | 14 | Describe any methods used to assess risk of bias due to missing results in a synthesis (arising from reporting biases). | Page 7-8 |
| **Certainty assessment** | 15 | Describe any methods used to assess certainty (or confidence) in the body of evidence for an outcome. | Page 7-8 |
| **RESULTS** |  |  |  |
| **Study selection** | 16a | Describe the results of the search and selection process, from the number of records identified in the search to the number of studies included in the review, ideally using a flow diagram. | Page 8 |
|  | 16b | Cite studies that might appear to meet the inclusion criteria, but which were excluded, and explain why they were excluded. | Page 8 |
| **Study characteristics** | 17 | Cite each included study and present its characteristics. | Page 8-9, Table 1 |
| **Risk of bias in studies** | 18 | Present assessments of risk of bias for each included study. | Page 8-9, Table 2, Appendix files |
| **Results of individual studies** | 19 | For all outcomes, present, for each study: (a) summary statistics for each group (where appropriate) and (b) an effect estimate and its precision (e.g. confidence/credible interval), ideally using structured tables or plots. | Page 9-10 |
| **Results of syntheses** | 20a | For each synthesis, briefly summarise the characteristics and risk of bias among contributing studies. | Page 9-10 |
|  | 20b | Present results of all statistical syntheses conducted. If meta-analysis was done, present for each the summary estimate and its precision (e.g. confidence/credible interval) and measures of statistical heterogeneity. If comparing groups, describe the direction of the effect. | Page 9-10 |
|  | 20c | Present results of all investigations of possible causes of heterogeneity among study results. | Page 9-10 |
|  | 20d | Present results of all sensitivity analyses conducted to assess the robustness of the synthesized results. | Page 9-10 |
| **Reporting biases** | 21 | Present assessments of risk of bias due to missing results (arising from reporting biases) for each synthesis assessed. | N/A |
| **Certainty of evidence** | 22 | Present assessments of certainty (or confidence) in the body of evidence for each outcome assessed. | Page 9-10, appendix files |
| **DISCUSSION** |  |  |  |
| **Discussion** | 23a | Provide a general interpretation of the results in the context of other evidence. | Page 10-13 |
|  | 23b | Discuss any limitations of the evidence included in the review. | Page 11-13 |
|  | 23c | Discuss any limitations of the review processes used. | Page 12-13 |
|  | 23d | Discuss implications of the results for practice, policy, and future research. | Page 13 |
| **OTHER INFORMATION** |  |  |  |
| **Registration and protocol** | 24a | Provide registration information for the review, including register name and registration number, or state that the review was not registered. | Page 5 |
|  | 24b | Indicate where the review protocol can be accessed, or state that a protocol was not prepared. | Page 5 |
|  | 24c | Describe and explain any amendments to information provided at registration or in the protocol. | Page 5 |
| **Support** | 25 | Describe sources of financial or non-financial support for the review, and the role of the funders or sponsors in the review. | Title page |
| **Competing interests** | 26 | Declare any competing interests of review authors. | Title page |
| **Availability of data, code and other materials** | 27 | Report which of the following are publicly available and where they can be found: template data collection forms; data extracted from included studies; data used for all analyses; analytic code; any other materials used in the review. | N/A |

**PRIMSA Abstract Checklist**

| **Topic** | **No.** | **Item** | **Reported?** |
| --- | --- | --- | --- |
| **TITLE** |  |  |  |
| **Title** | 1 | Identify the report as a systematic review. | Yes |
| **BACKGROUND** |  |  |  |
| **Objectives** | 2 | Provide an explicit statement of the main objective(s) or question(s) the review addresses. | Yes |
| **METHODS** |  |  |  |
| **Eligibility criteria** | 3 | Specify the inclusion and exclusion criteria for the review. | Yes |
| **Information sources** | 4 | Specify the information sources (e.g. databases, registers) used to identify studies and the date when each was last searched. | No |
| **Risk of bias** | 5 | Specify the methods used to assess risk of bias in the included studies. | No |
| **Synthesis of results** | 6 | Specify the methods used to present and synthesize results. | No |
| **RESULTS** |  |  |  |
| **Included studies** | 7 | Give the total number of included studies and participants and summarise relevant characteristics of studies. | Yes |
| **Synthesis of results** | 8 | Present results for main outcomes, preferably indicating the number of included studies and participants for each. If meta-analysis was done, report the summary estimate and confidence/credible interval. If comparing groups, indicate the direction of the effect (i.e. which group is favoured). | Yes |
| **DISCUSSION** |  |  |  |
| **Limitations of evidence** | 9 | Provide a brief summary of the limitations of the evidence included in the review (e.g. study risk of bias, inconsistency and imprecision). | Yes |
| **Interpretation** | 10 | Provide a general interpretation of the results and important implications. | Yes |
| **OTHER** |  |  |  |
| **Funding** | 11 | Specify the primary source of funding for the review. | No |
| **Registration** | 12 | Provide the register name and registration number. | Yes |

*From:* Page MJ, McKenzie JE, Bossuyt PM, Boutron I, Hoffmann TC, Mulrow CD, et al. The PRISMA 2020 statement: an updated guideline for reporting systematic reviews. MetaArXiv. 2020, September 14. DOI: 10.31222/osf.io/v7gm2. For more information, visit: <www.prisma-statement.org>
